# Association of reduced estimated GFR with mortality in men and women across the adult age spectrum in the Optum Labs Data Warehouse

**DOI:** 10.1101/2025.03.27.25324539

**Authors:** Josef Coresh, Yingying Sang, Shoshana H. Ballew, Aditya Surapaneni, Morgan E. Grams

**Author notes:** **Address for Correspondence:** Chronic Kidney Disease Prognosis Consortium (Dr. Josef Coresh), 227 East 30th, room 707, New York, NY 10003.

## Abstract

Chronic kidney disease (CKD) is associated with multiple adverse outcomes. This study quantifies the mortality risk associated with CKD, characterized by an estimated glomerular filtration rate (eGFR) of <60 ml/min/1.73m^2^, utilizing de-identified electronic health record data from the Optum Labs Data Warehouse in 4,788,021 men and 5,766,551 women. Mortality rates were estimated per 1000 person-years by sex and 5-year age group and absolute risk difference were estimated as attributable risk per 1000 person-years. Elevated mortality rates were seen among individuals with reduced eGFR for all age groups and for both men and women. The analysis revealed a linear decline in the incidence rate ratios of mortality with advancing age, while attributable risks increased due to the marked increase in mortality with age. These sex-specific risk estimates from a large sample enhance previous findings and are crucial for refining global burden of disease metrics and health economic evaluations of CKD.

## Brief Report

Chronic kidney disease (CKD) is associated with an increased risk of mortality as well as other adverse outcomes.^1^ The presence of kidney damage (e.g., albuminuria), or an estimated glomerular filtration rate (eGFR) threshold of <60 ml/min/1.73m^2^ is used to define CKD.^2^ This eGFR threshold has been widely used to estimate the burden of kidney dysfunction and the associated excess risk of mortality, including by the Global Burden of Disease study.^3^ At older ages, the relative risk of mortality decreases while attributable risks associated with eGFR increase.^4^ The extent to which this happens similarly in men and women is less clear and useful for updating global burden of disease and health economic analyses.

This study used de-identified electronic health record (EHR) data from the Optum Labs Data Warehouse (OLDW). The database contains longitudinal health information on enrollees and patients, representing a mixture of ages and geographical regions across the United States. The EHR-derived data includes a subset of EHR data that has been normalized and standardized into a single database.^5^ The OLDW was used to define a baseline population with measured baseline serum creatinine (after 2012 and at least one year after enrollment in the system). Estimated GFR (eGFR) was calculated using the race-free CKD-EPI equation^6^ and dichotomized at 60 ml/min/1.73m^2^, with values above 105 ml/min/1.73m^2^ excluded to provide a more uniform comparison group, since high eGFR values form the J-shaped part of the risk relationship.^1^ Mortality and censoring were defined using the deceased indicator from hospital discharge and last encounter data, respectively. Mortality rates were estimated per 1000 person-years by sex and 5-year age group. Their ratios and differences, along with standard errors and 95% confidence intervals, were calculated using the normal approximation.^7^ Mortality rates were graphed vs. age group and sex on the log scale to demonstrate multiplicative interaction. Absolute risk difference (attributable risk per 1000 person years) was graphed on the arithmetic scale to demonstrate additive interaction. Formal tests of interaction were not conducted since the emphasis was on estimates and precision for clinical significance rather than statistical significance.

Mortality rates were higher among patients with eGFR <60 ml/min/1.73m^2^ compared to those above this threshold in all age groups for both men and women (**Table 1**). The incidence rate ratios of mortality associated with lower eGFR decreased with age linearly in both men and women, but all estimates and 95% confidence intervals were greater than 1 (**Figure 1**). The attributable risks to lower eGFR increased with age due to the marked increase in mortality with age (**Figure 2**).

**Table 1.**
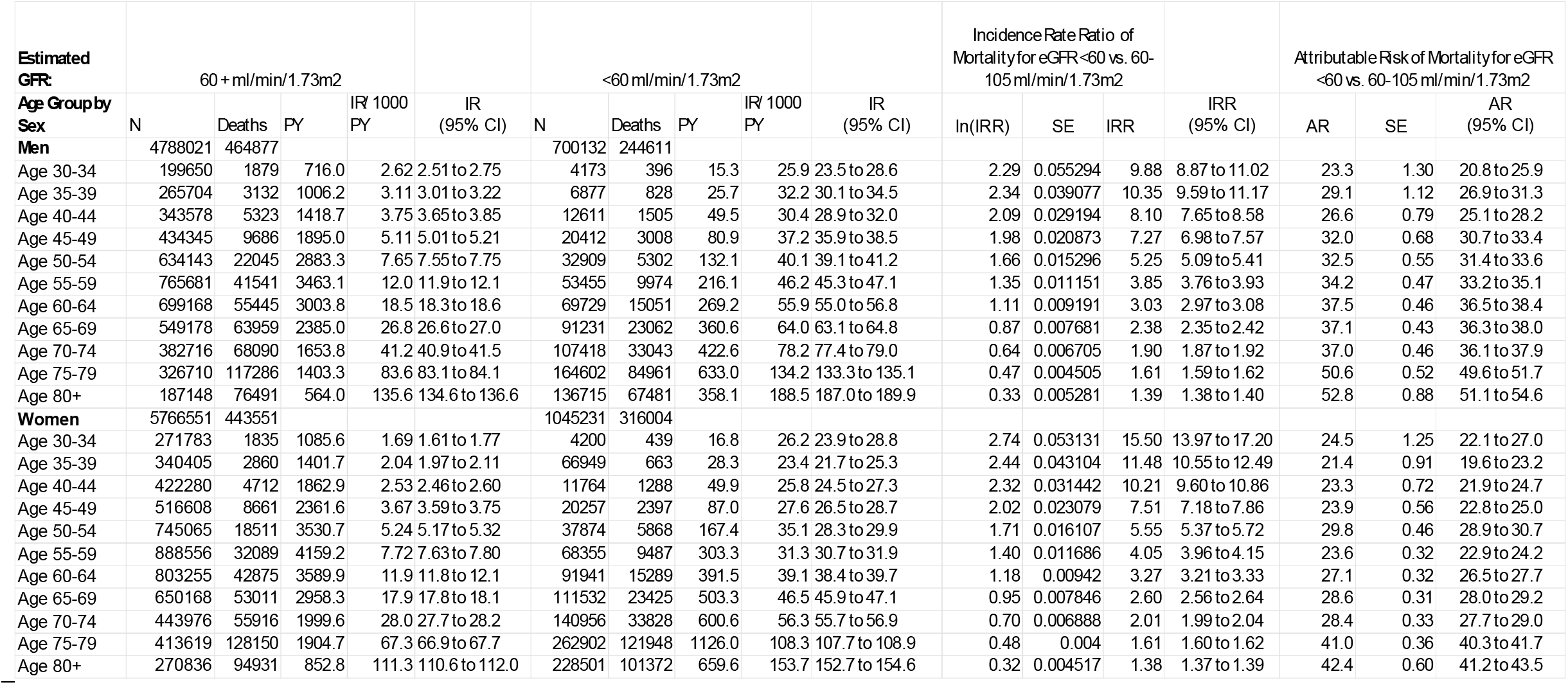
Mortality rates associated with low estimated GFR (<60 ml/min/1.73 m^2^) compared to higher estimated GFR (60-115 ml/min/1.73 m^2^) by age and sex among adult patients in the Optum Labs Data Warehouse.

| Estimated GFR: | 60 + ml/min/1.73m2 |  |  |  |  | <60 ml/min/1.73m2 |  |  |  |  | Incidence Rate Ratio of Mortality for eGFR <60 vs. 60-105 ml/min/1.73m2 |  |  | Attributable Risk of Mortality for eGFR <60 vs. 60-105 ml/min/1.73m2 |  |  |  |
| --- | --- | --- | --- | --- | --- | --- | --- | --- | --- | --- | --- | --- | --- | --- | --- | --- | --- |
| Age Group by Sex | N | Deaths | PY | IR/ 1000 PY | IR (95% CI) | N | Deaths | PY | IR/ 1000 PY | IR (95% CI) | ln(IRR) | SE | IRR | IRR (95% CI) | AR | SE | AR (95% CI) |
| Men | 4788021 | 464877 |  |  |  | 700132 | 244611 |  |  |  |  |  |  |  |  |  |  |
| Age 30-34 | 199650 | 1879 | 716.0 | 2.62 | 2.51 to 2.75 | 4173 | 396 | 15.3 | 25.9 | 23.5 to 28.6 | 2.29 | 0.055294 | 9.88 | 8.87 to 11.02 | 23.3 | 1.30 | 20.8 to 25.9 |
| Age 35-39 | 265704 | 3132 | 1006.2 | 3.11 | 3.01 to 3.22 | 6877 | 828 | 25.7 | 32.2 | 30.1 to 34.5 | 2.34 | 0.039077 | 10.35 | 9.59 to 11.17 | 29.1 | 1.12 | 26.9 to 31.3 |
| Age 40-44 | 343578 | 5323 | 1418.7 | 3.75 | 3.65 to 3.85 | 12611 | 1505 | 49.5 | 30.4 | 28.9 to 32.0 | 2.09 | 0.029194 | 8.10 | 7.65 to 8.58 | 26.6 | 0.79 | 25.1 to 28.2 |
| Age 45-49 | 434345 | 9686 | 1895.0 | 5.11 | 5.01 to 5.21 | 20412 | 3008 | 80.9 | 37.2 | 35.9 to 38.5 | 1.98 | 0.020873 | 7.27 | 6.98 to 7.57 | 32.0 | 0.68 | 30.7 to 33.4 |
| Age 50-54 | 634143 | 22045 | 2883.3 | 7.65 | 7.55 to 7.75 | 32909 | 5302 | 132.1 | 40.1 | 39.1 to 41.2 | 1.66 | 0.015296 | 5.25 | 5.09 to 5.41 | 32.5 | 0.55 | 31.4 to 33.6 |
| Age 55-59 | 765681 | 41541 | 3463.1 | 12.0 | 11.9 to 12.1 | 53455 | 9974 | 216.1 | 46.2 | 45.3 to 47.1 | 1.35 | 0.011151 | 3.85 | 3.76 to 3.93 | 34.2 | 0.47 | 33.2 to 35.1 |
| Age 60-64 | 699168 | 55445 | 3003.8 | 18.5 | 18.3 to 18.6 | 69729 | 15051 | 269.2 | 55.9 | 55.0 to 56.8 | 1.11 | 0.009191 | 3.03 | 2.97 to 3.08 | 37.5 | 0.46 | 36.5 to 38.4 |
| Age 65-69 | 549178 | 63959 | 2385.0 | 26.8 | 26.6 to 27.0 | 91231 | 23062 | 360.6 | 64.0 | 63.1 to 64.8 | 0.87 | 0.007681 | 2.38 | 2.35 to 2.42 | 37.1 | 0.43 | 36.3 to 38.0 |
| Age 70-74 | 382716 | 68090 | 1653.8 | 41.2 | 40.9 to 41.5 | 107418 | 33043 | 422.6 | 78.2 | 77.4 to 79.0 | 0.64 | 0.006705 | 1.90 | 1.87 to 1.92 | 37.0 | 0.46 | 36.1 to 37.9 |
| Age 75-79 | 326710 | 117286 | 1403.3 | 83.6 | 83.1 to 84.1 | 164602 | 84961 | 633.0 | 134.2 | 133.3 to 135.1 | 0.47 | 0.004505 | 1.61 | 1.59 to 1.62 | 50.6 | 0.52 | 49.6 to 51.7 |
| Age 80+ | 187148 | 76491 | 564.0 | 135.6 | 134.6 to 136.6 | 136715 | 67481 | 358.1 | 188.5 | 187.0 to 189.9 | 0.33 | 0.005281 | 1.39 | 1.38 to 1.40 | 52.8 | 0.88 | 51.1 to 54.6 |
| Women | 5766551 | 443551 |  |  |  | 1045231 | 316004 |  |  |  |  |  |  |  |  |  |  |
| Age 30-34 | 271783 | 1835 | 1085.6 | 1.69 | 1.61 to 1.77 | 4200 | 439 | 16.8 | 26.2 | 23.9 to 28.8 | 2.74 | 0.053131 | 15.50 | 13.97 to 17.20 | 24.5 | 1.25 | 22.1 to 27.0 |
| Age 35-39 | 340405 | 2860 | 1401.7 | 2.04 | 1.97 to 2.11 | 66949 | 663 | 28.3 | 23.4 | 21.7 to 25.3 | 2.44 | 0.043104 | 11.48 | 10.55 to 12.49 | 21.4 | 0.91 | 19.6 to 23.2 |
| Age 40-44 | 422280 | 4712 | 1862.9 | 2.53 | 2.46 to 2.60 | 11764 | 1288 | 49.9 | 25.8 | 24.5 to 27.3 | 2.32 | 0.031442 | 10.21 | 9.60 to 10.86 | 23.3 | 0.72 | 21.9 to 24.7 |
| Age 45-49 | 516608 | 8661 | 2361.6 | 3.67 | 3.59 to 3.75 | 20257 | 2397 | 87.0 | 27.6 | 26.5 to 28.7 | 2.02 | 0.023079 | 7.51 | 7.18 to 7.86 | 23.9 | 0.56 | 22.8 to 25.0 |
| Age 50-54 | 745065 | 18511 | 3530.7 | 5.24 | 5.17 to 5.32 | 37874 | 5868 | 167.4 | 35.1 | 28.3 to 29.9 | 1.71 | 0.016107 | 5.55 | 5.37 to 5.72 | 29.8 | 0.46 | 28.9 to 30.7 |
| Age 55-59 | 888556 | 32089 | 4159.2 | 7.72 | 7.63 to 7.80 | 68355 | 9487 | 303.3 | 31.3 | 30.7 to 31.9 | 1.40 | 0.011686 | 4.05 | 3.96 to 4.15 | 23.6 | 0.32 | 22.9 to 24.2 |
| Age 60-64 | 803255 | 42875 | 3589.9 | 11.9 | 11.8 to 12.1 | 91941 | 15289 | 391.5 | 39.1 | 38.4 to 39.7 | 1.18 | 0.00942 | 3.27 | 3.21 to 3.33 | 27.1 | 0.32 | 26.5 to 27.7 |
| Age 65-69 | 650168 | 53011 | 2958.3 | 17.9 | 17.8 to 18.1 | 111532 | 23425 | 503.3 | 46.5 | 45.9 to 47.1 | 0.95 | 0.007846 | 2.60 | 2.56 to 2.64 | 28.6 | 0.31 | 28.0 to 29.2 |
| Age 70-74 | 443976 | 55916 | 1999.6 | 28.0 | 27.7 to 28.2 | 140956 | 33828 | 600.6 | 56.3 | 55.7 to 56.9 | 0.70 | 0.006888 | 2.01 | 1.99 to 2.04 | 28.4 | 0.33 | 27.7 to 29.0 |
| Age 75-79 | 413619 | 128150 | 1904.7 | 67.3 | 66.9 to 67.7 | 262902 | 121948 | 1126.0 | 108.3 | 107.7 to 108.9 | 0.48 | 0.004 | 1.61 | 1.60 to 1.62 | 41.0 | 0.36 | 40.3 to 41.7 |
| Age 80+ | 270836 | 94931 | 852.8 | 111.3 | 110.6 to 112.0 | 228501 | 101372 | 659.6 | 153.7 | 152.7 to 154.6 | 0.32 | 0.004517 | 1.38 | 1.37 to 1.39 | 42.4 | 0.60 | 41.2 to 43.5 |

**Figure 1.**
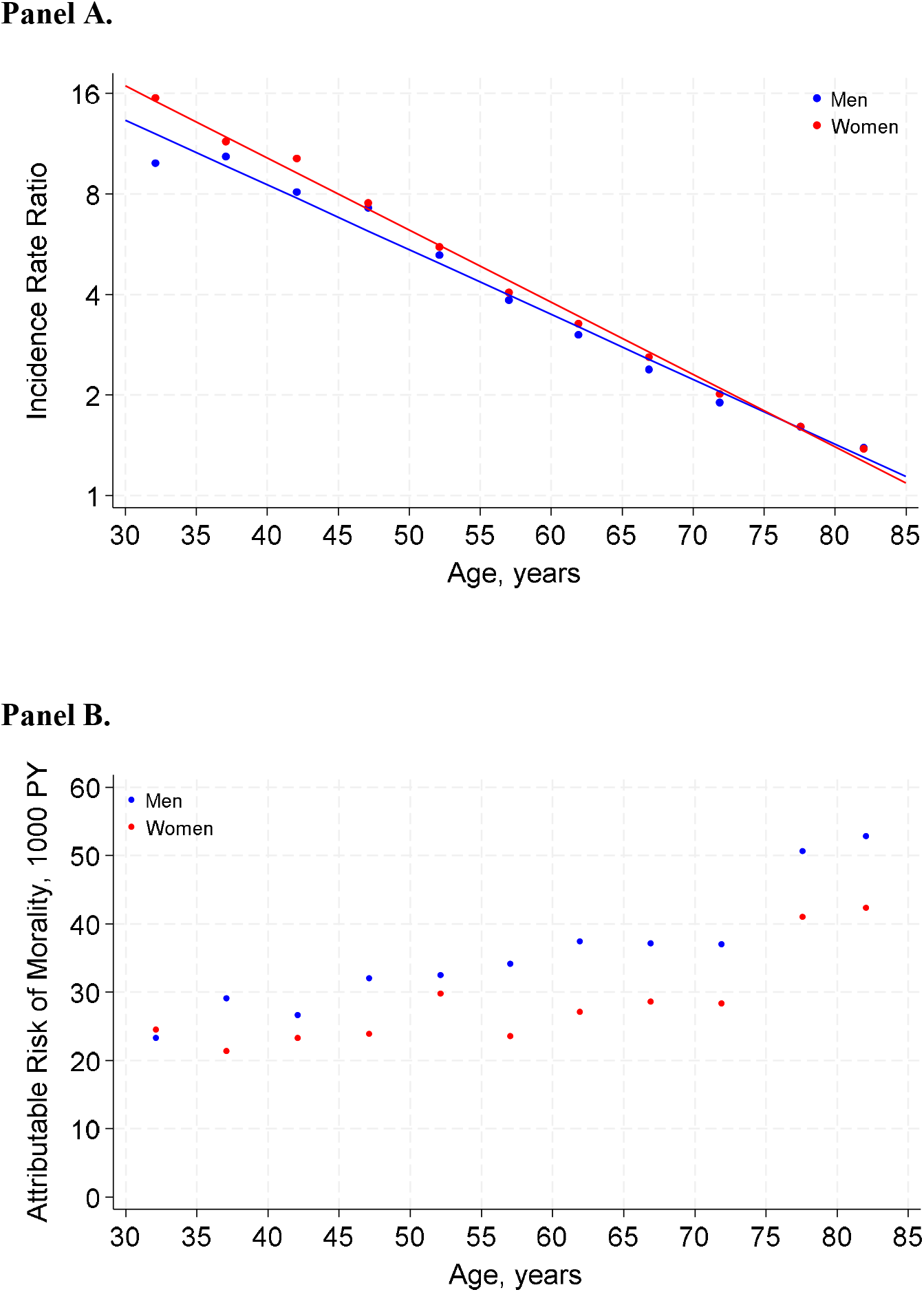
Risk of mortality associated with low estimated GFR (<60 ml/min/1.73 m^2^) compared to higher estimated GFR (60-115 ml/min/1.73 m^2^) by age and sex displayed are incidence rate ratios (panel A) and attributable risk (panel B).

In summary, this paper quantifies the mortality risk associated with low eGFR across age and sex strata. This report provides sex-specific estimates of risk, relative risks, and attributable risks, extending previous studies that reported overall adjusted associations.^4^ Strengths of the study include the large sample size and precise estimates. Limitations include the lack of adjustment beyond age and sex and the dichotomization of kidney function, although results are consistent with the adjusted continuous analysis in Hallan et al.^4^ These data and similar meta-analyzed relative risks can be used in updating global burden of disease estimates for the risk and disability-adjusted life years (DALYs) associated with kidney dysfunction,^3^ as well as health economic analyses of CKD burden.

## Data Availability

Under agreement with the participating cohorts, CKD-PC cannot share individual data with third parties. Inquiries regarding specific analyses should be made to.

## Sources of Funding

The CKD Prognosis Consortium (CKD-PC) Data Coordinating Center is funded in part by a program grant from the US National Kidney Foundation and the National Institute of Diabetes and Digestive and Kidney Diseases (R01DK100446).

## Disclosures

None

## Data Sharing Statement

Under agreement with the OLDW we cannot share individual data with third parties. Inquiries regarding specific analyses should be made to.

## Notes

### Competing Interest Statement

The authors have declared no competing interest.

### Author Declarations

The institutional review board at the New York University Grossman School of Medicine, New York, NY, USA (#i23-00919) gave ethical approval for this work.

## REFERENCES

1. Grams ME, Coresh J, Matsushita K, et al. Estimated Glomerular Filtration Rate, Albuminuria, and Adverse Outcomes: An Individual-Participant Data Meta-Analysis. JAMA. Oct 3 2023;330(13):1266–1277. doi:10.1001/jama.2023.17002

2. Kidney Disease: Improving Global Outcomes CKD Work Group. KDIGO 2024 Clinical Practice Guideline for the Evaluation and Management of Chronic Kidney Disease. Kidney Int. Apr 2024;105(4S):S117–S314. doi:10.1016/j.kint.2023.10.018

3. GBD Chronic Kidney Disease Collaboration. Global, regional, and national burden of chronic kidney disease, 1990-2017: a systematic analysis for the Global Burden of Disease Study 2017. Lancet. Feb 29 2020;395(10225):709–733. doi:10.1016/S0140-6736(20)30045-3

4. Hallan SI, Matsushita K, Sang Y, et al. Age and Association of Kidney Measures With Mortality and End-stage Renal Disease. JAMA. Oct 30 2012;308(22):2349–2360. doi:10.1001/jama.2012.168171387683 [pii]

5. Optum Labs. Optum Labs and Optum Labs Data Warehouse (OLDW) Descriptions and Citation. Vol. PDF. Reproduced with permission from Optum Labs. PDF. March 2023.

6. Inker LA, Eneanya ND, Coresh J, et al. New Creatinine- and Cystatin C-Based Equations to Estimate GFR without Race. N Engl J Med. Nov 4 2021;385(19):1737–1749. doi:10.1056/NEJMoa2102953

7. StataCorp. Stata Base Reference Manual Release 18. Stata Press; 2023.

